# Decontamination of N95 masks against coronavirus: a scoping review

**DOI:** 10.1101/2020.07.11.20151399

**Authors:** Rafael Sarkis-Onofre, Rafaela do Carmo Borges, Giulia Demarco, Lara Dotto, Falk Schwendicke, Flávio Fernando Demarco

## Abstract

**Background:** At present, it remains uncertain which method to decontaminate N95 is most suitable and should be recommended to healthcare professionals worldwide.

**Objectives:** The aim of this scoping review was to map and compile the available evidence about the effectiveness of decontaminating N95 masks against coronavirus.

**Methods:** We selected studies written in English assessing or discussing decontamination strategies of N95 masks against coronavirus. The search and study screening were performed in PubMed and SCOPUS by two independent researchers. A descriptive analysis was performed considering the study design of included studies.

**Results:** We included nineteen studies. Eight articles were letter to the editors, five were in vitro studies, three were literature reviews, and three were classified as other study designs. The use of vaporized hydrogen peroxide and ultraviolet irradiation were the strategies most cited. However, there is a lack of evidence and consensus related to the best method of N95 masks decontamination.

**Conclusion:** The evidence towards decontamination strategies of N95 masks against coronavirus remains scarce. Vaporized hydrogen peroxide and ultraviolet irradiation seem the current standard for N95 masks decontamination.

## Introduction

The novel Coronavirus, known as SARS-Cov2, has produced a social disruption globally, with severe consequences for general health of the population. At the present moment, there are more than 11 million confirmed cases, with a cumulative number of deaths of over 500,000 according to World Health Organization (WHO) (updated data can be accessed in: https://www.worldometers.info/coronavirus/). Currently, vaccines are still under trial and there are no effective drugs for the treatment of this disease^1^. Indeed, most of the available evidence provided is that social distancing, wearing masks and eye protection are effective in the prevention of transmission^2^. Also, better hygiene (hand washing) and use of sanitizers are supported to detain spreading of the disease^3-5^.

The use of masks has been recommended by WHO, and around the world governments have established face protection policies in public space^6^. The resulting increase in demand and a shortage in their availability in the market have led to price spikes for masks^7-11^. Health professionals are at high risk for infection with the new coronavirus, and a lack of adequate protective equipment during critical procedures in infected patients is increasing that risk considerably^12, 13^. In Brazil, for example, more nurses and nurses’ assistants have died due to Covid-19 than anywhere else^14^ and most of them have been infected during their work with infected patients.

N95 masks are a type of respirator mask used as a facial protection specifically by healthcare providers^9, 15^. They have the capacity to filter over 95% of pollutant particles (>0.3 µm) in the air and have been suggested to be used to reduce the risk of Covid-19 spreading, too^16^. Due to their high costs and limited availability^8, 15^, different methods to decontaminate N95 masks^7, 15, 17-21^ has been discussed to allow multiple usage.

Decontamination methods can be classified into chemical or radiation treatment, dry heat or moist heat^9^. Such methods need to fulfill certain criteria: elimination of all pathogens; no damage to the facemask structure; the filter capacity of masks should stay the same; and no residue of the decontamination process should remain^9, 15^. At present, it remains uncertain which method to decontaminate N95 is most suitable and should be recommended to healthcare professionals worldwide. The aim of this scoping review was to map and compile the evidences about the effectiveness of different decontamination strategies of N95 masks against coronavirus.

## Methods

The protocol of this study is based on the framework proposed by Peters et al., 2015^22^ and is available at the following link: https://osf.io/4t936/. The reporting of this scoping review was based on PRISMA Extension for Scoping Reviews^23^.

### Eligibility criteria

We selected studies assessing different decontamination strategies of N95 masks against coronavirus or discussing decontamination strategies such as letters, editorials and literature review. No specifications towards the coronavirus organisms (surrogate or not) used to test decontamination or the decontamination strategies themselves were applied.

### Information sources and search

The search was performed in two databases: Medline (PubMed) and Scopus; only articles written in English language were included. The search strategy was based on MeSH terms of PubMed and specific terms of Scopus and the last search was performed in May 2020.

The following strategies were used:

#### PubMed

(((“Decontamination” [Mesh] OR “Decontamination” OR “Disinfection” OR “Ultraviolet-C” OR “peracetic acid”)) AND (“Masks” [Mesh] OR “Masks” OR “Respiratory Protective Devices” [Mesh] OR “Respiratory Protective Devices” OR “Device, Respiratory Protective” OR “Devices, Respiratory Protective” OR “Protective Device, Respiratory” OR “Protective Devices, Respiratory” OR “Respiratory Protective Device” OR “Respirators, Industrial” OR “Industrial Respirators” OR “Industrial Respirator” OR “Respirator, Industrial” OR “Respirators, Air-Purifying” OR “Air-Purifying Respirator” OR “Air-Purifying Respirators” OR “Respirator, Air-Purifying” OR “Respirators, Air Purifying” OR “N95”)) AND (“SARS-CoV-2” OR “Coronavirus” OR “COVID-19” OR “Coronaviruses”)

#### SCOPUS

“Decontamination” OR “Disinfection” OR “Ultraviolet-C” OR “peracetic acid” AND “Masks” OR “Respiratory Protective Devices” OR “Device, Respiratory Protective” OR “Devices, Respiratory Protective” OR “Protective Device, Respiratory” OR “Protective Devices, Respiratory” OR “Respiratory Protective Device” OR “Respirators, Industrial” OR “Industrial Respirators” OR “Industrial Respirator” OR “Respirator, Industrial” OR “Respirators, Air-Purifying” OR “Air-Purifying Respirator” OR “Air-Purifying Respirators” OR “Respirator, Air-Purifying” OR “Respirators, Air Purifying” OR “N95” AND “SARS-CoV-2” OR “Coronavirus” OR “COVID-19” OR “Coronaviruses”

### Selection

The search was undertaken using EndNote (EndNote X9, Thomson Reuters, New York, US). Two researches independently identified relevant records by analyzing titles and abstracts for relevance according to the eligibility criteria. Retrieved records were classified as include, exclude, or uncertain. The full-text articles of the included and uncertain records were selected for further eligibility screening by the same two reviewers, again independently. Discrepancies in screening of titles/abstracts and full text articles were resolved through a discussion. In case of disagreement, the opinion of a third reviewer was garnered.

### Data charting and items

We created a form using Excel (Microsoft, Redmond, Washington, US), which was pilot tested by three reviewers to reach a consensus on what data to collect and how. Then, two reviewers extracted the data independently, this was reviewed by a third reviewer. The following data were collected: study design; study objective, decontamination regimens tested, organisms studied, method of evaluation and main findings. For studies only discussing (and not reporting on) decontamination strategies, the following data were collected: study design, strategies discussed and main findings.

### Synthesis

A descriptive analysis was performed considering the study design and decontamination regimens tested or discussed.

## Results

The literature search yielded 178 titles and abstracts (Fig. 1). Nineteen studies^7, 8, 15, 17-21, 24-34^ fulfilled the eligibility criteria from which the data were extracted. Reasons for exclusion are listed in the Supplemental Material.

**Figure 1:**
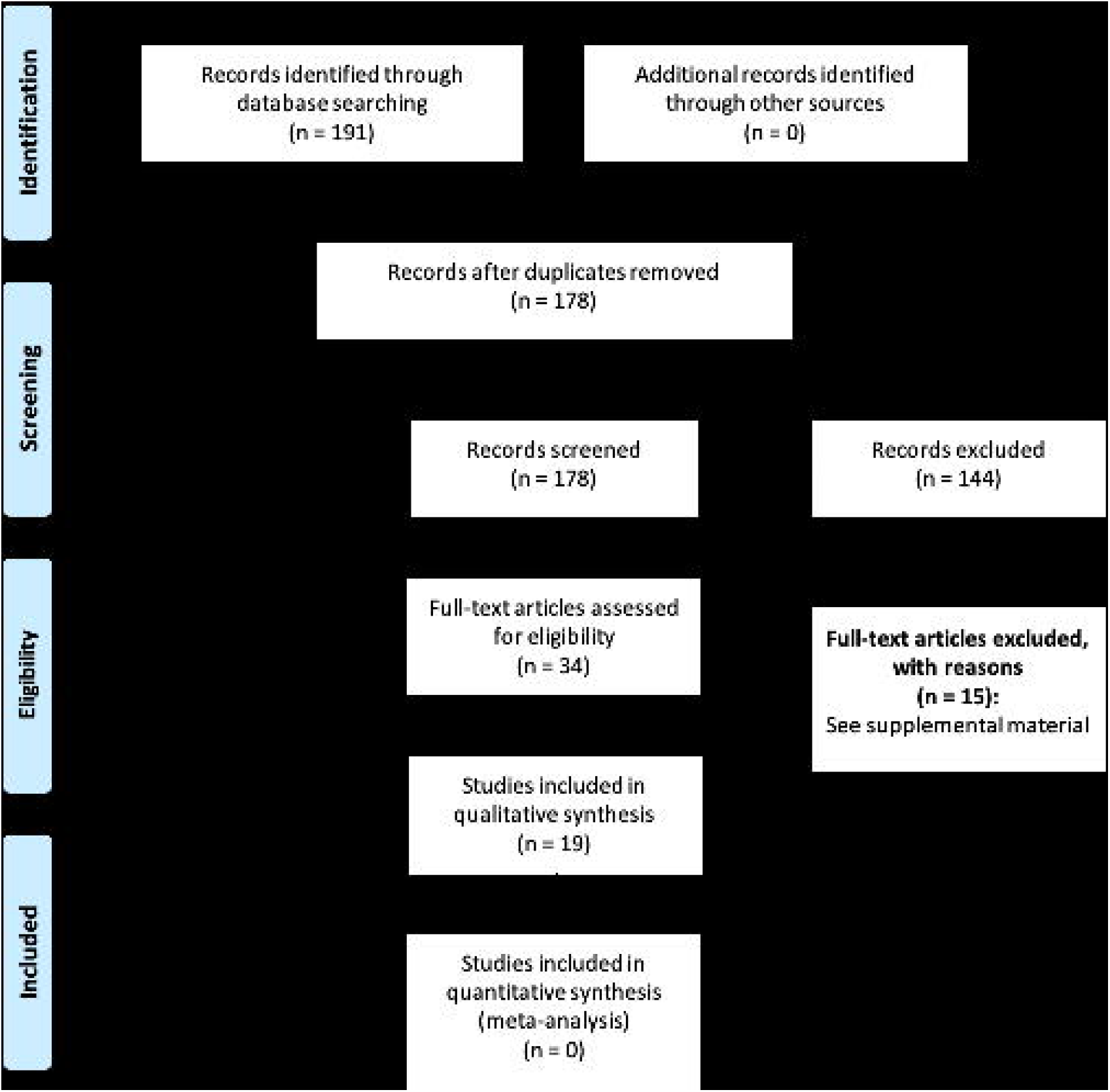
Flowchart of identification, screening, and assessing studies for inclusion eligibility

### Characteristics of included studies

Table I presents the characteristics of included studies. Related to study design of included studies, eight articles were letter to the editors^8, 15, 18, 20, 26, 28, 29, 33^, five were in vitro studies ^7, 19, 21, 25, 31^, three were literature reviews ^17, 24, 27^, and three were classified as other study designs ^30, 32, 34^. Considering only the eight letters to the editors, three letters discussed results of in vitro studies^8, 15, 18^.

**Table I:**
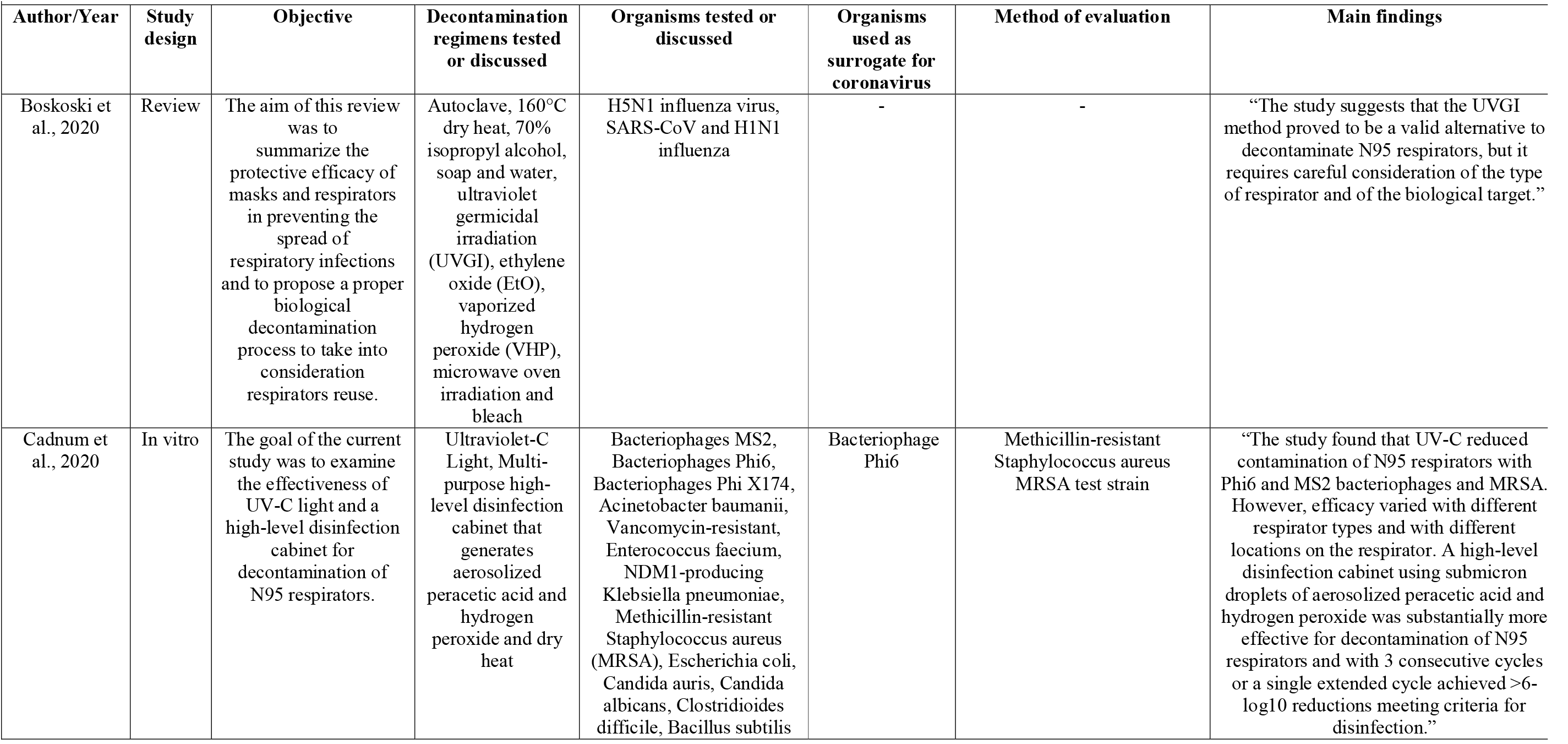
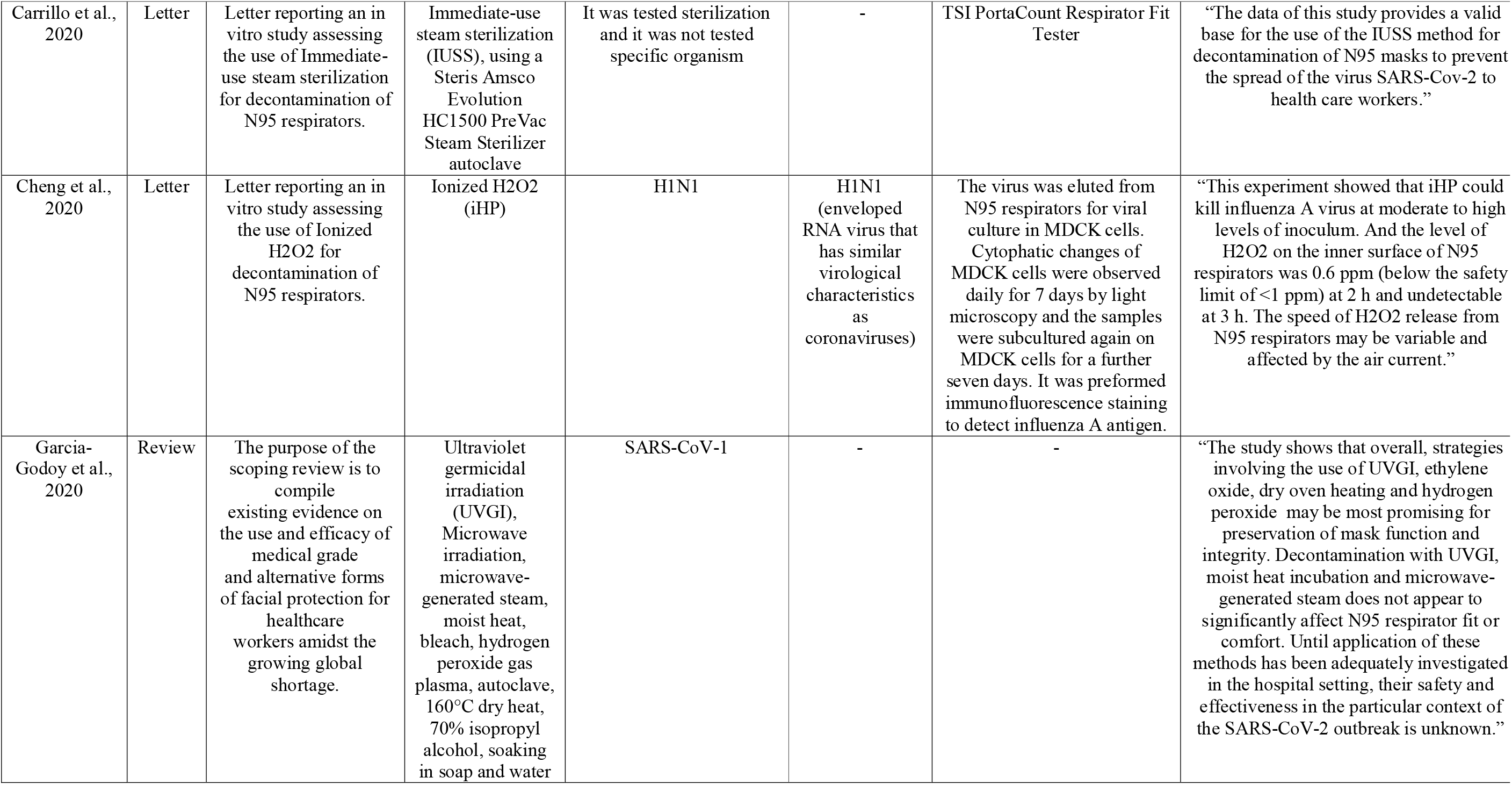
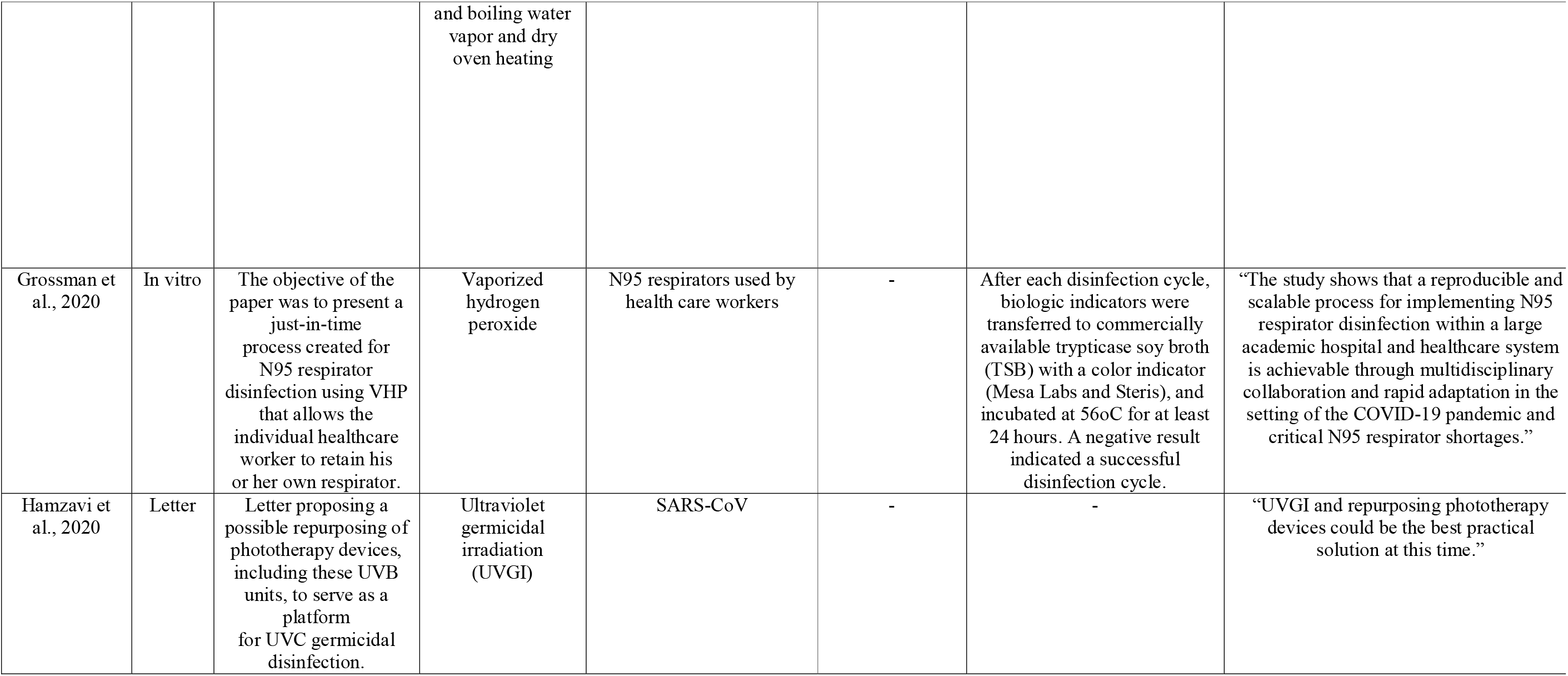
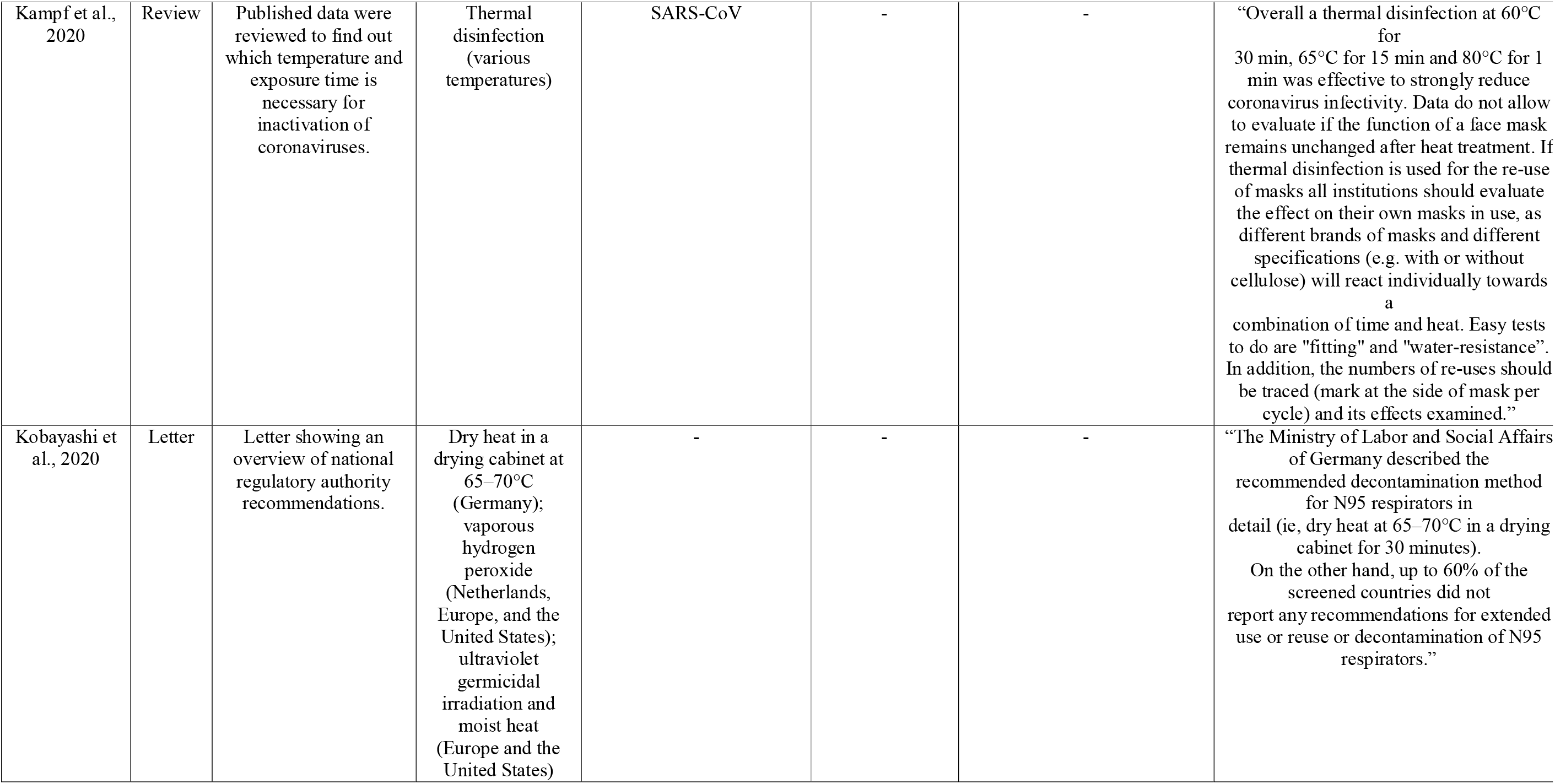
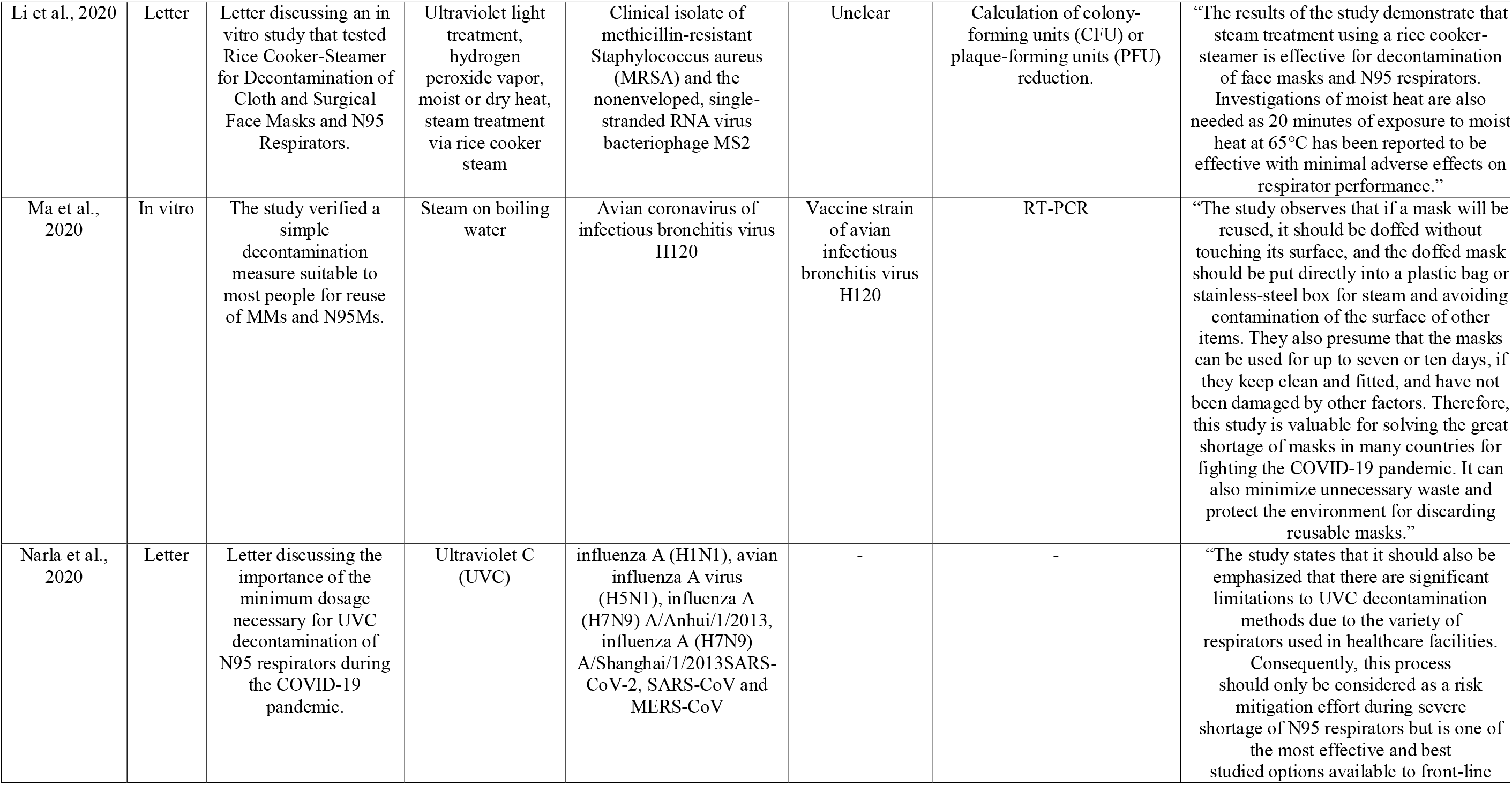
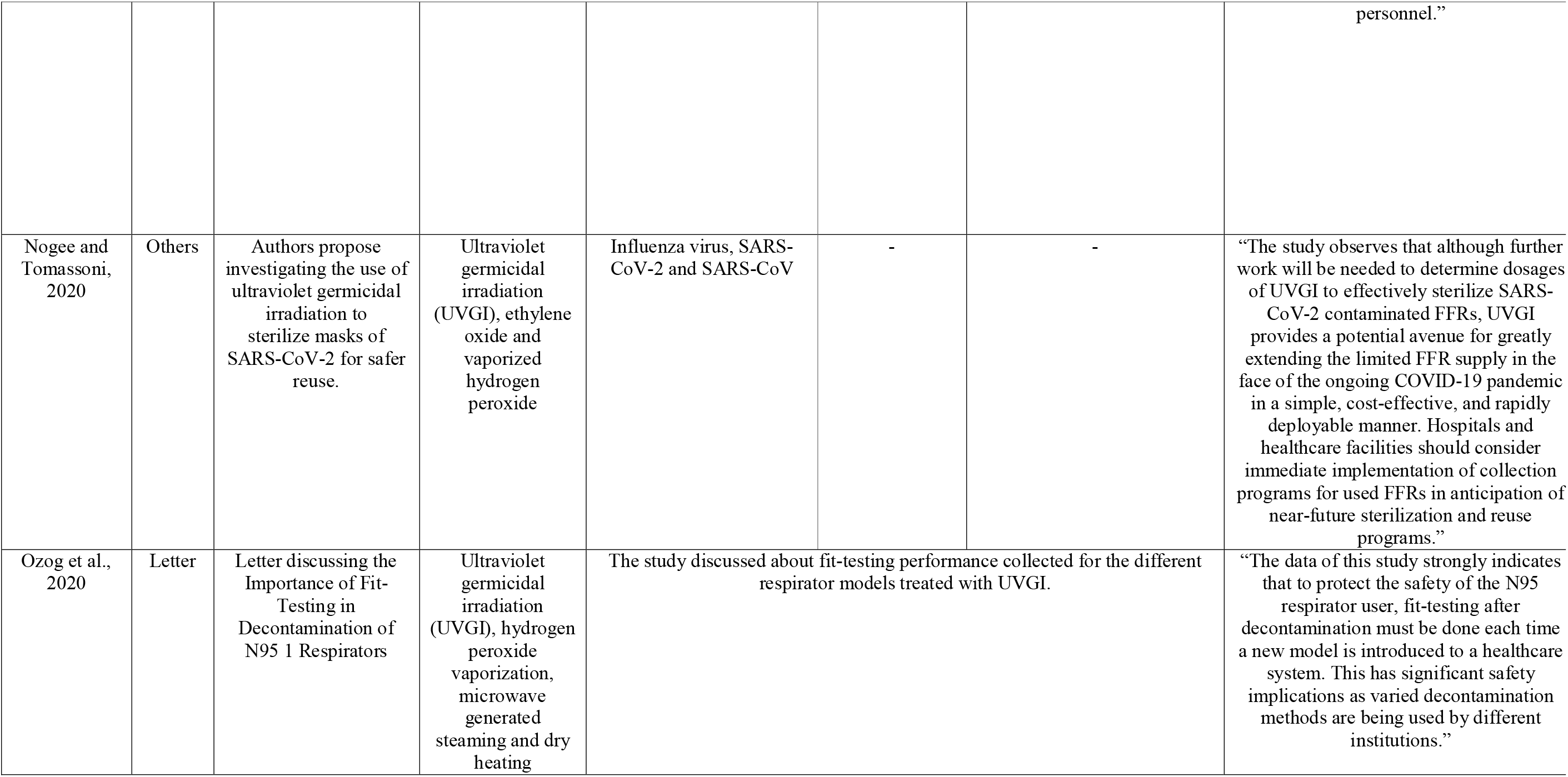
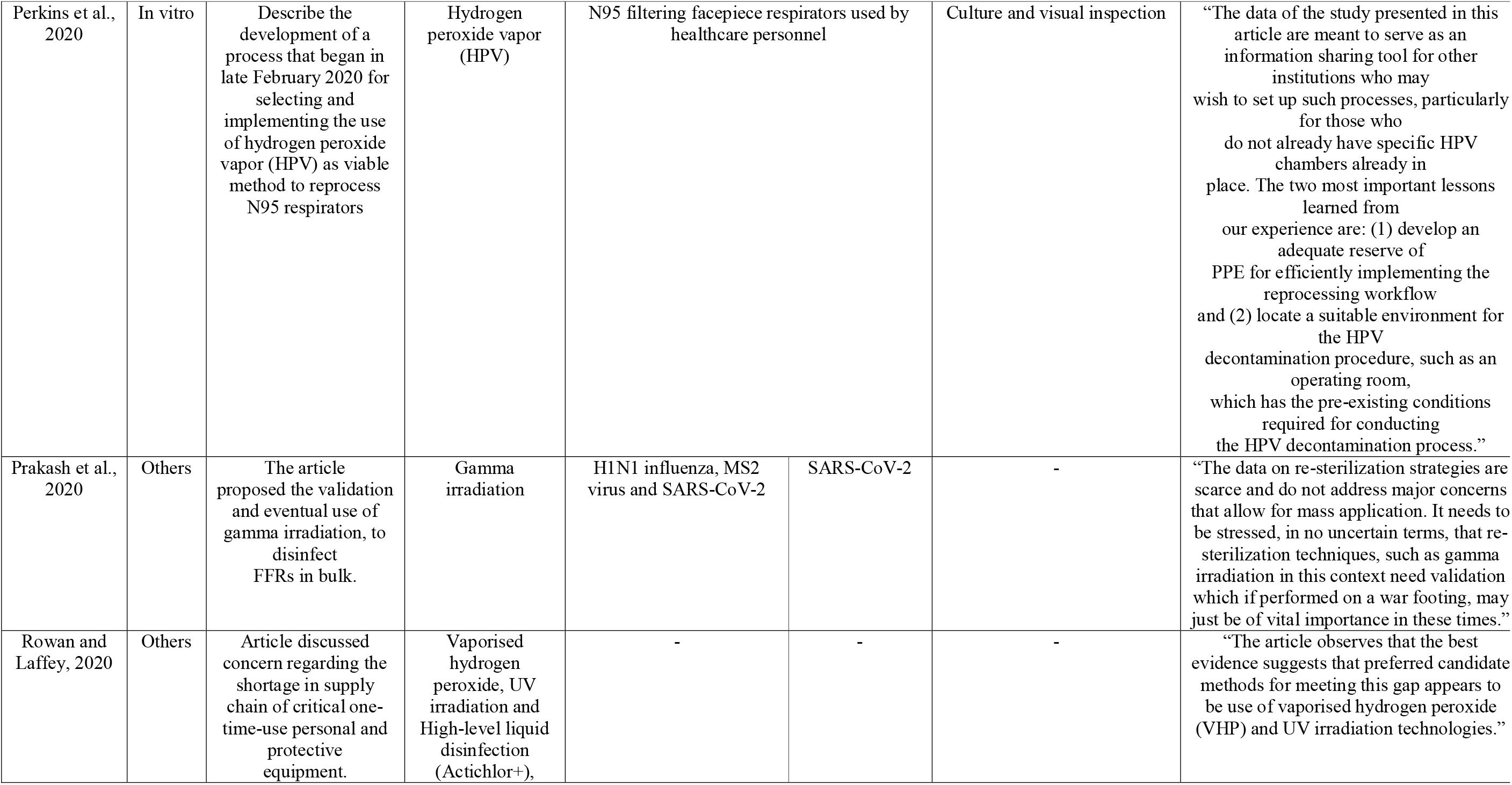
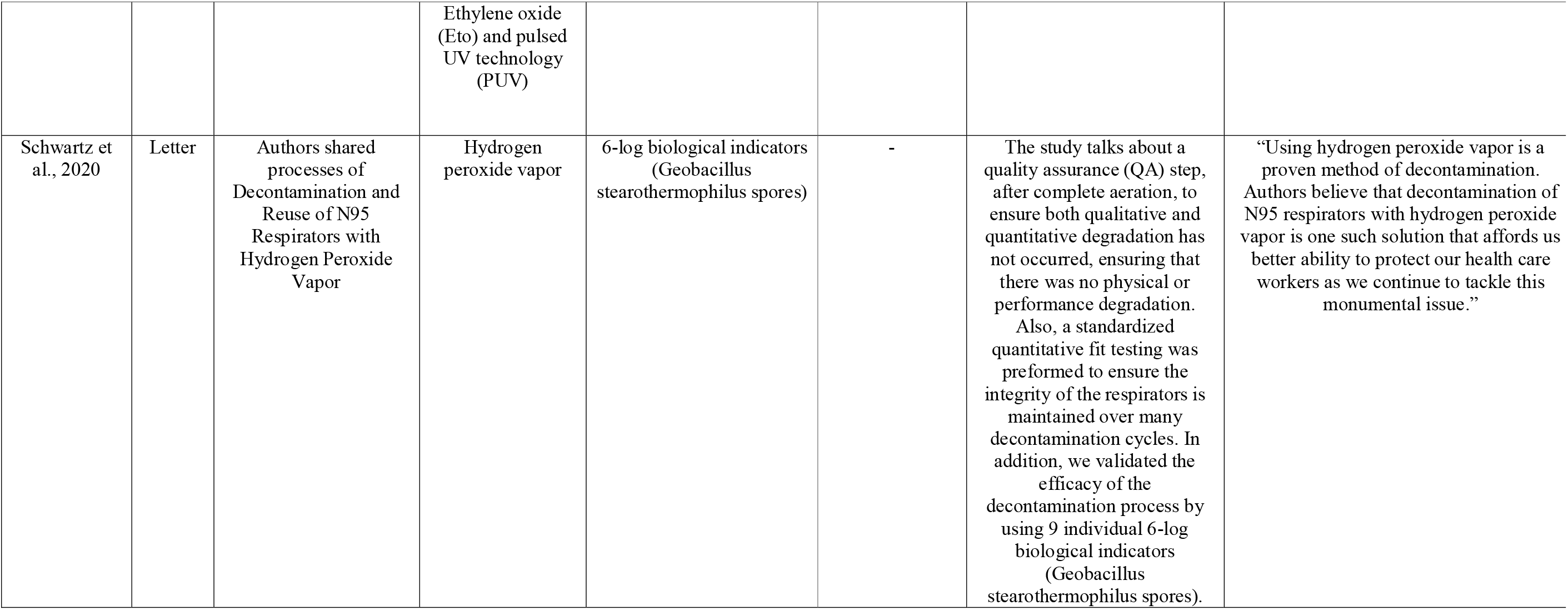
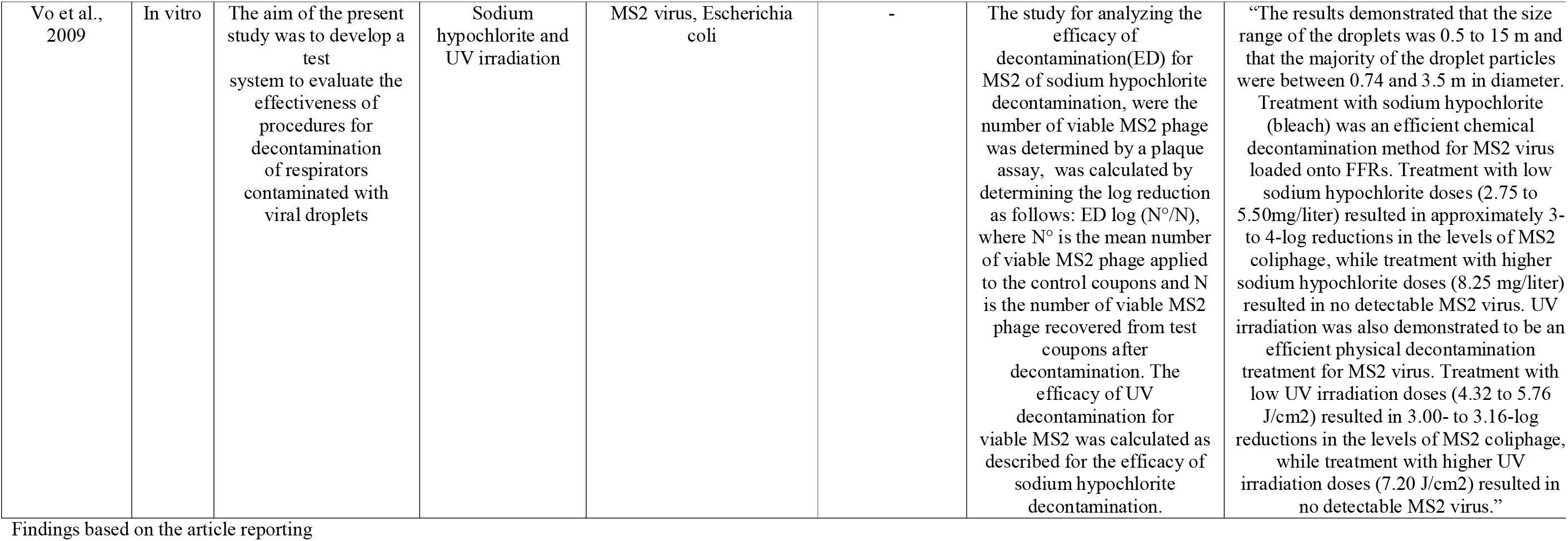
Characteristics of included studies

Related to decontamination regimens tested or discussed, the use of vaporized hydrogen peroxide and ultraviolet irradiation were the regimens most cited. The use of vaporized hydrogen peroxide was cited in four letters^18, 20, 28, 33^, three in vitro studies ^7, 25, 31^, two reviews^17, 24^ and two other study designs^30, 32^. The use of ultraviolet irradiation vaporized hydrogen peroxide was cited in five letters^9, 20, 26, 28, 29^, two in vitro studies^7, 21^, two reviews^17, 24^ and two other study designs ^30, 32^.

### Vaporized hydrogen peroxide

Three studies reported the process for N95 decontamination with vaporized hydrogen peroxide. Schwartz et al. 2020 described the process implemented in the Duke University (US) and demonstrated that vaporized hydrogen peroxide is an efficacious decontamination method that does not cause physical or performance degradation of the masks^33^.

Perkins et al. 2020 presented the process implemented in the University of New Mexico (US) and reported on the low toxicity of the methods. The authors highlighted the importance of physical assessment of the mask after decontamination^31^.

Grossman et al. 2020 described the decontamination using vaporized hydroperoxide employed by Washington University (US). They demonstrated that the entire process requires less than 24 hours and showed that it is important to create a workflow to achieve an effective decontamination considering pre-processing steps, decontamination process and, post-processing steps^25^.

Further studies evaluated that strategy combined with others or discussed its availability and feasibility. Cadnum et al. 2020 performed an in vitro study and compared the use of a high-level decontamination cabinet that generates aerosolized peracetic acid and hydrogen peroxide with Ultraviolet-C light and dry heat at 70^0^C for 30 minutes. They demonstrated that aerosolized peracetic acid and hydrogen peroxide are effective for decontamination of N95^7^.

Kobayashi et al. 2020 assessed the authority recommendations in the Netherlands, the states governments in the US, and the European Commission Directorate-General for Health and Consumers as well as the European Medicines Agency on the use of vaporous hydrogen peroxide. They found that while this method seems to lead to acceptable decontamination while retaining mask integrity according to visual inspection, this type of decontamination is not available throughout all countries and institutions and currently no standard for its application exists^28^.

Garcia-Godoy et al. 2020 and Rowan et al. 2020 discussed that the use of vaporous hydrogen peroxide seems to be one of the most promising method for N95 decontamination^24, 32^.

### Ultraviolet C light

Hamzanzi et al., 2020 presented a prototype model for N95 decontamination using Ultraviolet germicidal irradiation that would allow decontamination of 18 to 27 masks in one process^26^.

Kobayashi et al. 2020 assessed the authority recommendations on the use of Ultraviolet germicidal irradiation and found that while this method is promising, it has not been standardized by any of the authorities so far^28^.

Cadnum et al. 2020 demonstrated that the use of Ultraviolet C could reduce N95 contamination, but efficacy varied with different masks types and locations on the respirator^7^.

Vo et al. 2009 showed that high doses of Ultraviolet irradiation (>7.20 J/cm2; UV intensity, 0.4 mW/cm2; contact times, >5 h) could inactivate virus loaded in N95 masks^21^.

Garcia-Godoy et al. 2020 discussed that the Ultraviolet germicidal irradiation seems to be one of the most promising method for N95 decontamination^24^.

Narla et al. 2020 highlighted that is necessary at least 1 J/cm2 to all surfaces to ensure N95 decontamination. However, the authors emphasized that Ultraviolet decontamination has limitations, mainly as each type of mask needs a specific dosage of irradiation to be reliably effective^29^.

### Other methods

Carillo et al. 2020 indicated that immediate use steam sterilization is an applicable decontamination method^15^.

Li et al. 2020 demonstrated that steam treatment using a rice cooker-steamer is effective for decontamination of N95 masks^18^.

Vo et al 2009, showed that treatment with sodium hypochlorite was an effective decontamination method^21^.

Ma et al. 2020 demonstrated that steam decontamination could be used as a decontamination of N95 masks^19^.

Kampf et al. 2020 showed that a thermal decontamination at 60°C for 30 min, 65°C for 15 min and 80°C for 1 min was effective to reduce coronavirus infectivity^27^.

## Discussion

This is the first study to map the evidence about the effectiveness of decontamination strategies of N95 masks against coronavirus. Our results demonstrate that there is a lack of evidence and consensus related to the best method of N95 masks decontamination. However, the use of vaporized hydrogen peroxide and ultraviolet irradiation were the regimens most cited and seem to be the most promising methods for N95 masks decontamination.

Hydrogen peroxide vapor decontamination is a common method used in different fields and facilities, including scientific, pharmaceutical, and medical ones. The method has low toxicity and uses the catalytic reduction of peroxide to oxygen and water^35^. However, it needs a specific room and equipment to achieve an effective decontamination and, hence, is rather expensive. Ultraviolet irradiation is a method of decontamination using Ultraviolet light to inactivate microorganisms through DNA damage and jeopardizing cell functions^36^. The use of this decontamination method has limitations due different mask needing different irradiation dosages; high dosage in turn could result in high toxicity and structural damage of the mask. Moreover, it also needs specific equipment, limiting its availability.

Ideally, any decontamination method should eliminate all pathogens; maintain mask integrity and filter capacity; at low toxicity and costs. Until a method does not fulfill these criteria, the extended use of masks seems to be a good and low-cost approach to overcome the discussed limitations in availability. Current recommendations consider mask wearing periods between 4 and 40 hours ^28^. Notably, additional protection such as use of face shield and strict adherence to hand hygiene practices are needed, especially if extending mask wearing periods^37^.

Outcomes such as mechanical integrity and performance of N95 masks should be observed when assessing decontamination strategies of N95 masks, as decontamination may come at a price; decontaminated but not effective masks are not useful or even dangerous. Ozog et al., 2020 indicated that a fit testing must be performed after decontamination and if a decontamination is achieved but the masks lost their integrity, further usage should be stopped^20^. Hence, both integrity and performance should be prioritized when implementing decontamination strategies, while not all included studies concomitantly tested decontamination and subsequent performance of the mask.

Our study presents some limitations. First, because this was a scoping review, we did not conduct a risk of bias/quality assessment of the included studies. Second, we included only studies in English. Third, the included studies present different designs and protocols, making it difficult to compare the results specially because many brands of N95 are available on the market, different regimens were tested and individual scenarios of wearers (such as, influence of as cosmetics or sunscreens use for Ultraviolet decontamination) are difficult to test. Finally, we included article discussing decontamination methods based on opinions over evidences making it difficult better conclusions and recommendations.

Considering that the global pandemic accelerates its spread at present, and taking into account the shortage of protective equipment, especially for healthcare workers, more investigations for safely decontaminating N95 masks are needed. Also, the availability and cost-effectiveness of decontamination should be considered by future studies.

## Conclusion

The evidence towards decontamination strategies of N95 masks against coronavirus remains scarce. Vaporized hydrogen peroxide and ultraviolet irradiation seem the current standard for N95 masks decontamination.

## Data Availability

The authors confirm that the data supporting the findings of this study are available within the article and its supplementary materials.

## Acknowledgments

RSO is funded in part by Meridional Foundation (Passo Fundo, Brazil) and LD is funded by Coordination for the Improvement of Higher Education Personnel (CAPES, Brazil). RCB is funded by National Council for Scientific and Technological Development (CNPQ, Brazil) and FFD is funded in part by National Council for Scientific and Technological Development (CNPQ, Brazil).

## Contribution

RSO − Conceptualization, Methodology, Investigation, Project administration, Supervision, Writing-original draft.

RCB - Data curation, Investigation, Writing – Review & Editing

GD - Data curation, Investigation, Writing – Review & Editing

LD - Data curation, Investigation, Writing – Review & Editing

FS - Methodology, Validation, Writing-review & editing.

FFD - Conceptualization, Methodology, Project Administration, Writing – Review & Editing, Funding acquisition.

